# UNIVERSAL EPIDEMIC SCALING: INFLUENZA AND COVID-19

**DOI:** 10.64898/2026.09.16.26363217

**Authors:** Denis Below, Felix Mairanowski

## Abstract

Conventional compartmental models may become difficult to parameterize over multi-wave epidemic horizons when susceptibility and transmission conditions change between successive epidemic regimes. This study introduces a reduced macroscopic framework that changes the scale of epidemic description from individual-level transmission structure to effective population-level dynamics. Epidemic waves are formulated as transport processes governed by deterministic boundaries, mass balance, and a small set of effective macroscopic parameters. The susceptible population is represented by a dynamic Effective Susceptible Pool that can be re-initialized at transitions between biologically distinct epidemic regimes, while transmission resistance and external control measures are incorporated at the macroscopic level. The resulting equations admit a dimensionless similarity representation and closed-form analytical solutions, enabling analytical estimation of epidemic trajectories and peak healthcare demand without computationally intensive numerical simulation. The framework is evaluated using comparative time-series data for SARS-CoV-2 and seasonal influenza A within the geographically and demographically consistent setting of Rhode Island. Despite their different biological and immunological regimes, the analyzed trajectories exhibit a common reduced asymptotic scaling form. The results support the use of a macroscopic, scale-reduced representation for cross-calibration of heterogeneous surveillance signals and analytical assessment of healthcare-system demand. Further validation across pathogens, populations, and open-system settings is required.

**Highlights:**

- Epidemic waves are modeled analytically as macroscopic transport processes in non-linear media.
- The vulnerable population is redefined as a dynamic, re-initialized effective susceptible pool.
- Mass immunization acts as a synchronized control operator suppressing the epidemic peak.
- A strict mass-balance deconvolution filter removes post-symptomatic shedding noise.
- Peak hospital capacity thresholds are predicted using an analytically tractable closed-form method.

## 1. Introduction: Macroscopic System Foundations

Mathematical modeling of infectious disease transmission faces an epistemological tension between granular, compartment-based parameterization (e.g., SEIR-class numerical frameworks) and macroscopic, non-linear system aggregation. While microscale compartmental models are valuable for isolated, short-term epidemic episodes, their deployment over multi-wave horizons—particularly for rapidly mutating RNA viruses—can lead to substantial non-stationarity in parameterization, parameter cross-correlation, and over-parameterization. The COVID-19 pandemic, especially the abrupt emergence of antigenically distinct lineages, highlighted these critical limitations: models calibrated primarily on cumulative population immunity may fail to capture the phase-space transformation, velocity, and magnitude of new variant-driven surges

In the spirit of Plutarch’s Parallel Lives, which sought to extract universal principles of human character by comparing distinct historical figures, this study proposes an alternative macro-evolutionary similarity framework. Rather than treating pandemic emergencies and routine endemic seasonal oscillations as distinct topological phenomena, we analyze the parallel trajectories of two epidemiologically contrasting agents—an emergent pandemic pathogen (SARS-CoV-2 ancestral strain) and an endemic agent (seasonal influenza A)—within a single, demographically controlled domain: the State of Rhode Island. This "Plutarchian" comparative matrix isolates intrinsic dynamical variations from demographic, environmental, and behavioral baselines, allowing us to uncover scale-invariant universal laws governing both systems.

Our approach revives the macroscopic tradition initiated by Daniel Bernoulli [2,3] and formalized by Kermack and McKendrick **[1]**, treating epidemic waves as deterministic transport processes governed by mass balance rather than microscale compartmental chaos [4]. From the perspective of mathematical physics, an epidemic wave functions as a self-limiting trajectory within a non-linear medium. Here, the intrinsic transmission coefficient k_0_ operates as the macroscopic driving force of the viral flux, while the behavioral contact reduction parameter λ acts as an internal negative feedback coefficient, representing the social system’s dynamic resistance to perceived infection risk. Concurrently, external biological and technological constraints, such as progressive viral attenuation and mass immunization, act as control operators that systematically truncate the effective boundary of the Effective Susceptible Pool S(t) defined here as a macroscopic measure of transmission-accessible population capacity rather than literal biological susceptibility.

By applying dimensional analysis and the Buckingham Pi-theorem [5,6] to this coupled system, the parameter space is reduced to a minimal set of dimensionless similarity groups, a methodology comprehensively developed in the classical treatise by Sedov[6]. Under this transformation, the evolutionary timeline of an independent wave collapses onto a scale-invariant universal trajectory whose normalized form is independent of the individual dimensional parameter values and is governed by asymmetric Gompertz kinetics [7,8]. The applicability of Gompertz-class sigmoidal growth functions to viral spreading has been extensively validated in recent literature [9].

Building upon recent multi-country simulations [10, 11] and previous frameworks on urban confinement constraints, this paper transitions the paradigm from empirical curve-fitting to a universal deterministic engine. We demonstrate that the global evolution of independent epidemic waves, when re-initialized at each variant boundary, is governed by invariant non-linear criteria that can be evaluated via closed-form analytical expressions. Crucially, we incorporate established observations on antigenic drift, immune escape, and tissue-tropism shifts to interpret how highly transmissible pandemic surges can transition toward stable, self-limiting endemic regimes analogous, at the macroscopic dynamical level, to seasonal influenza.

## 2. Conceptual Foundations of Macro-Evolutionary Epidemic Wave Dynamics

Before formalizing the mathematical apparatus, we outline the macroscopic principles that motivate the proposed framework. These principles establish a conceptual bridge between statistical physics, non-linear dynamic systems, and evolutionary virology, challenging the continuous-immunity approximations embedded in classical compartmental structures [4,10–12].

### 2.1. Phenomenological Regimes: Endemic Baseline vs. Dissipative Surges

For established respiratory pathogens, a characteristic endemic baseline represents a steady-state transmission level sustained by a continuously replenished pool of susceptible individuals (via birth cohorts and waned immunity) and partial antigenic mismatch. In contrast to this established endemic regime, a novel pathogen entering an immunologically naive population operates in a highly non-equilibrium regime. In this setting, the pre-existing endemic baseline for the novel pathogen is effectively zero, while the initial effective susceptible pool is taken as S_0_=100% in the limiting case of an immunologically naïve population. This initial immunological vacuum generates a primary epidemic wave functioning as a macroscopic transport process in a non-linear, dissipative medium without pre-existing pathogen-specific population immunity. The phase-space structure of this primary surge is fundamentally distinct from routine endemic oscillations.

### 2.2. Antigenic Non-Stationarity and Evolutionary Attenuation as Drivers of System Reset

RNA viruses leverage high mutation rates to introduce structural non-stationarity into the population matrix, challenging the assumption that the effective susceptible pool can be represented solely as a continuous function of cumulative exposure. In our framework, we identify three distinct evolutionary mechanisms that reshape the boundary conditions of the transmission medium:

• **Antigenic Drift:** The gradual accumulation of point mutations in surface glycoproteins (e.g., influenza hemagglutinin [HA]) continuously modifies the boundary conditions of the transmission medium. Minor genetic shifts at critical antigenic sites systematically degrade the historical resistance of the population, reducing seasonal vaccine efficacy and driving recurring wave profiles [4,27,30].

• **Antigenic Shift:** Segment reassortment introduces novel viral subtypes that effectively bypass the historically accumulated immune memory of the host population, instantaneously re-initializing the transmission capacity of the network [4].

**Coronaviral Immune Escape and Pathological Attenuation:** While genetic reassortment is absent in SARS-CoV-2, rapid spike-protein mutations can produce a macroscopically analogous dynamic reset. However, the observed evolutionary trajectory of SARS-CoV-2 has also been associated with changes in the balance between transmission efficiency and clinical severity [13–16].

### 2.3. Coronaviral Escape and Phase-Space Resets

While genetic reassortment (antigenic shift) is absent in SARS-CoV-2, rapid spike-protein mutations can produce a macroscopically analogous dynamic reset. The emergence of the Omicron variant, carrying over 30 amino acid alterations in the spike glycoprotein, demonstrated that a virus can rapidly evade neutralizing antibodies, rendering past humoral immunity partially obsolete at the variant boundary.

In-vitro serological studies from this period confirm that neutralizing antibody (NAb) titers against new variants frequently dropped by factors of 10 to 40, occasionally falling below detectable thresholds. This profound immune evasion effectively resets the system’s boundary conditions, momentarily re-initializing the effective susceptible fraction and generating rapid, variant-driven surges [13–16].

Crucially, this dynamic is heavily influenced by host immune history. In both influenza and SARS-CoV-2, this imprinting acts as a directional constraint on the population’s immunological phase space [24–26]. Rather than generating a symmetrical immune response to the mutant pathogen, the host network’s immunological memory preferentially re-mobilizes antibodies toward ancestral, non-effective epitopes, complicating the direct translation of serological titers into sterilizing herd protection.

Within the proposed macroscopic framework, an epidemic wave cannot in general be represented solely as a continuous cumulative function of historical exposure when a substantial antigenic or immune escape transition occurs. Instead, a sufficiently large antigenic or immune escape transition is represented as a discrete macroscopic regime change, requiring a re-initialization of the effective susceptible boundary [27, 31].

### 2.4. Evolutionary Transition and Endemic Stabilization

A key requirement for a scale-invariant model is to explain how the non-equilibrium pandemic regime can transition toward a stable, background seasonal baseline—a phenomenon observed both in the final phase of the 1918 H1N1 influenza pandemic and in the post-pandemic phase of COVID-19.

Serological and clinical observations suggest that this stabilization is associated with an evolutionary shift in the balance between transmission efficiency and clinical severity. A reduction in clinical virulence may represent a recurrent tendency observed during prolonged viral evolution, although this tendency is neither universal nor monotonic and may be locally reversed by individual mutations or emerging lineages. In the case of SARS-CoV-2, late-stage Omicron lineages, including JN.1 and subsequent FLiRT-related lineages, were associated with altered tissue tropism. Reduced reliance on TMPRSS2-mediated cell entry and a greater propensity for replication in the upper respiratory tract have been associated with reduced pathogenicity and clinical severity relative to earlier lineages [22, 23, 32].

From an epidemiological standpoint, these lineages also exhibited substantial immune escape, resulting in markedly reduced antibody-mediated protection against infection. However, substantial cross-reactive T-cell memory, mediated by CD4+ and CD8+ T-cell responses, remained comparatively preserved despite extensive surface-antigenic change [33]. This preserved cellular immunity likely contributed to limiting severe disease, even as neutralizing-antibody protection against infection was substantially reduced.

At the macroscopic level, these evolutionary changes are consistent with a transition toward an endemic regime. Within each strain-specific wave, however, the attenuation of transmission is governed by the effective macroscopic resistance λ, rather than by progressive mutational attenuation of the ongoing strain. Within each strain-specific wave, transmission is progressively attenuated as the effective susceptible pool S(t) is depleted and the macroscopic resistance λ limits further propagation. This attenuation is therefore a property of the epidemic medium rather than a consequence of progressive mutational weakening of the ongoing strain.

Mutational and antigenic changes are instead treated as discrete transitions between strain-specific transmission regimes, capable of modifying the effective susceptible boundary S_0_, the transmission coefficient k, and, where biologically supported, the clinical severity characteristics of the pathogen. Thus, successive variants can re-initialize the effective susceptible pool S(t) while generating a new wave trajectory, whereas the subsequent attenuation of that wave is governed by the macroscopic resistance λ. Over successive strain transitions, the system can therefore move from an acute pandemic regime toward a stable endemic-like respiratory disease regime, with its normalized long-term trajectory converging toward the macroscopic behavior observed for seasonal influenza A.

## 3. Mathematical Formalism and Operational Postulates

The macroscopic principles established in Section 2 require a mathematical representation that departs from classical continuous-immunity frameworks. Rather than relying on continuous compartmental depletion, we formalize these dynamics into a decoupled state system capable of delivering closed-form, scale-invariant solutions.

### 3.1. System Field Equations

The primary field variable, S(t), functions as the Effective Susceptible Pool (or effective dynamic permeability) of the continuous social matrix. This variable tracks the instantaneous spatial and topological capacity of transmission pathways accessible to the viral flux. While the initial uncoupled prototype equations governing this baseline state space were introduced in [36], based on early threshold dynamics recorded in [37], the present study substantially extends the formalism into a complete similarity framework. To capture the simultaneous impact of internal behavioral friction and active technological interventions, the temporal evolution of the system is governed by the following coupled differential equations:

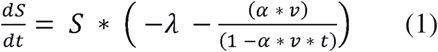

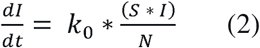

where k_0_ day^-1^ represents the intrinsic transmission rate (the macroscopic driving head of the viral stream), and λ day^-1^ is the macroscopic viscous drag coefficient representing dynamic behavioral friction (e.g., risk-induced compliance, contact reduction). The technological intervention layer is modeled as a linear control operator characterized by a uniform daily implementation velocity v day^-1^ and a containment efficiency factor α (denoting vaccine efficacy).

### 3.2. Mathematical Postulates and Boundaries of Validity

The analytical integrity and similarity scaling of the reduced system are defined by three foundational postulates:

### Postulate I: Autonomous Effective Susceptible Field

The Effective Susceptible Pool S(t) is treated as an autonomous macroscopic state variable describing the effective transmission permeability of the social matrix. Within each strain-specific wave segment, its evolution is governed by the macroscopic resistance and control processes defined by Eq. (1), rather than by an explicit feedback from cumulative infection or demographic depletion. The demographic population N is therefore treated as a stationary reference field, while variations in S(t) represent the dynamic modulation of effective transmission pathways.

### Postulate II: Piecewise-Continuous Resetting and Phase-Space Re-initialization

To model long-term, multi-wave evolutionary timelines without accumulating historical fitting errors, the global framework operates as a piecewise-continuous dynamical system. When a substantial antigenic or immune-escape transition alters the effective population susceptibility to a new viral lineage, the preceding wave is treated as a completed dynamical segment and the effective state is re-initialized at the corresponding transition boundary. The Effective Susceptible Pool is therefore assigned a new boundary condition for each subsequent strain-specific regime. The parameter set (S_0_, k_0_, λ) is updated discretely at transition nodes between viral lineages, represented mathematically by Heaviside-like step functions. This formulation preserves the local analytical structure of each wave while allowing the macroscopic state of the population to change between successive regimes.

### Postulate III: State Aggregation and Peak Logistical Response

When modeling the cumulative infection trajectory rather than active prevalence, the internal clinical recovery dynamics are not required to define the primary epidemic transport state. The clinical residence parameter γ therefore enters only in the downstream hospital-occupancy formulation. This state aggregation is consistent with the operational objective of the framework, which is to determine peak logistical thresholds and healthcare-system capacity requirements. Within a finite clinical environment, hospital occupancy represents a delayed response to the infection influx rather than a direct measure of instantaneous epidemic incidence. The hospital-occupancy profile can therefore be represented as a downstream integrative response to the infection trajectory, with γ retained explicitly in the corresponding clinical convolution or mass-balance formulation.

### 3.3. Closed-Form Solutions and Similarity Coordinates

Direct integration of the Effective Susceptible Pool field equation (Eq. 1) under an active immunization campaign yields the analytical trajectory:

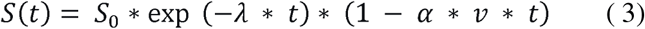

This product form separates the two macroscopic attenuation mechanisms: the exponential term represents the continuous contraction of effective transmission pathways associated with macroscopic resistance λ, whereas the linear term represents the time-dependent attenuation produced by the vaccination control αv.

For the baseline scenario without active vaccination, αv=0, the effective susceptible field reduces to

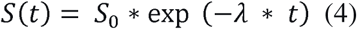

Substituting this total remaining Effective Susceptible path into the linearized transmission operator (Eq. 2) isolates a closed analytical trajectory for the infected fraction. In the baseline scenario prior to the deployment of vaccination (α * v = 0), the absolute system dynamics collapse onto an invariant coordinate system:

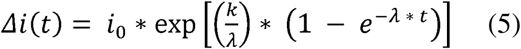

where Δi(t) = i(t) - i_f_, and i_f_ represents the cumulative infection fraction legacy inherited from the preceding wave at the temporal boundary where the new lineage emerges. The normalized cumulative infected population fraction, i(t), is defined as 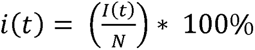

The macro-scale operational parameters are explicitly isolated as:

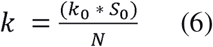

Here, i_0_ acts as the initial seed fraction anchoring the evolutionary impulse, standardized relative to the regional demographic matrix N as i_0_ = (100 / N)%, and k represents the effective macroscopic transmission coefficient. Mathematically, Eq. 6 takes the exact structural form of classical Gompertz growth kinetics. Its emergence in this macroscopic transmission system reflects the exponential accumulation of socio-behavioral resistance (λ) against the epidemic driving force, inherently limiting the outbreak wave.

Differentiation of the cumulative trajectory yields the incidence rate j(t)=di/dt. The maximum incidence is reached at:

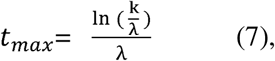

At this maximum-incidence point, the analytical solution gives a direct proportionality between the peak incidence j_max_ :

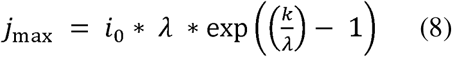

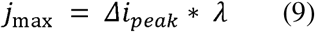

This isolates a fundamental, scale-invariant system asset: the behavioral contact reduction rate lambda is defined directly by the ratio of peak daily velocity to the accumulated wave volume (λ = /Δ ), completely decoupled from the absolute population size N.

Evaluating Eq. (5) at its long-term asymptotic boundary (t >∞) isolates the invariant logarithmic scaling ratio, which defines the total population fraction breached by the end of the unmitigated wave vector:

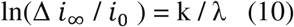

In diagnostic practice, i_∞_ is extracted directly from the terminal empirical plateau, providing a direct estimation of the dimensionless complex k / λ that is entirely decoupled from early-phase exponential noise. For ancestral viral vectors, the transmission parameter k was fixed by the geographic scale relationship:

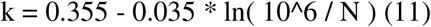

For emergent mutant lineages displaying enhanced transport kinetics, this scenario is systematically updated via the coupled mutation relationship:

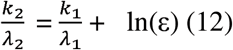

where ε represents the relative transmission advantage coefficient of the variant. Once k is fixed by the demographic domain, the corresponding λ is determined from the asymptotic invariant.

### 3.4. Initialization Diagnostics and Boundary Sensitivity Analysis

The mathematical transition from an invading pandemic vector to a deeply entrenched endemic regime requires a fundamental divergence in how the initial evolutionary seed fraction (i_0_) is configured. For an emergent pandemic expanding into an immunologically naive reservoir, the boundary condition can be modeled as a discrete, non-equilibrium impulse triggered by the introduction of the first clinical case into the region, where I_(0)_ = 1 case, yielding i_0_ = (100 / N)%.

Conversely, for an endemic agent such as Seasonal Influenza A, the pathogen is never fully cleared from the population matrix, smoldering subclinically through the summer months. To mirror this biological reality without introducing non-physical numerical singularities, our framework anchors the initial evolutionary impulse to the persistent interseasonal background incidence. At dynamic equilibrium, this background incidence coincides numerically with the initial incidence of the seasonal trajectory. For the present analysis, this boundary value is set to 50 cases per 1,000,000 population.

Crucially, this theoretical baseline is empirically validated by long-term regional surveillance. According to multi-year CDC ILINet tracking logs for New England, the absolute inter-seasonal outpatient registration floor during summer months operates within a stable corridor of approximately 0.5% to 1.0% of total healthcare traffic [34, 35]. Applying our inverse transport reconstruction framework to this clinical baseline isolates a real continuous circulation rate of approximately 65 true daily infections per million, demonstrating that our selected initialization boundary directly matches the lower physical limit of the target endemic ecosystem. To prove the structural robustness of the analytical Gompertz framework against the inherent uncertainty of this inter-seasonal background, a parameter sensitivity analysis was executed by varying the initial seed condition across three operational regimes: a high baseline (100 daily cases), a median baseline (50 daily cases), and a conservative low baseline (30 daily cases per million).

The resulting trajectory family exhibits high structural stability across these boundary shifts. Varying the initial equilibrium incidence from 30 to 100 cases per million population produces a systematic shift of the peak toward later times and a corresponding reduction in peak incidence as the initial boundary decreases.

**Figure 1.**
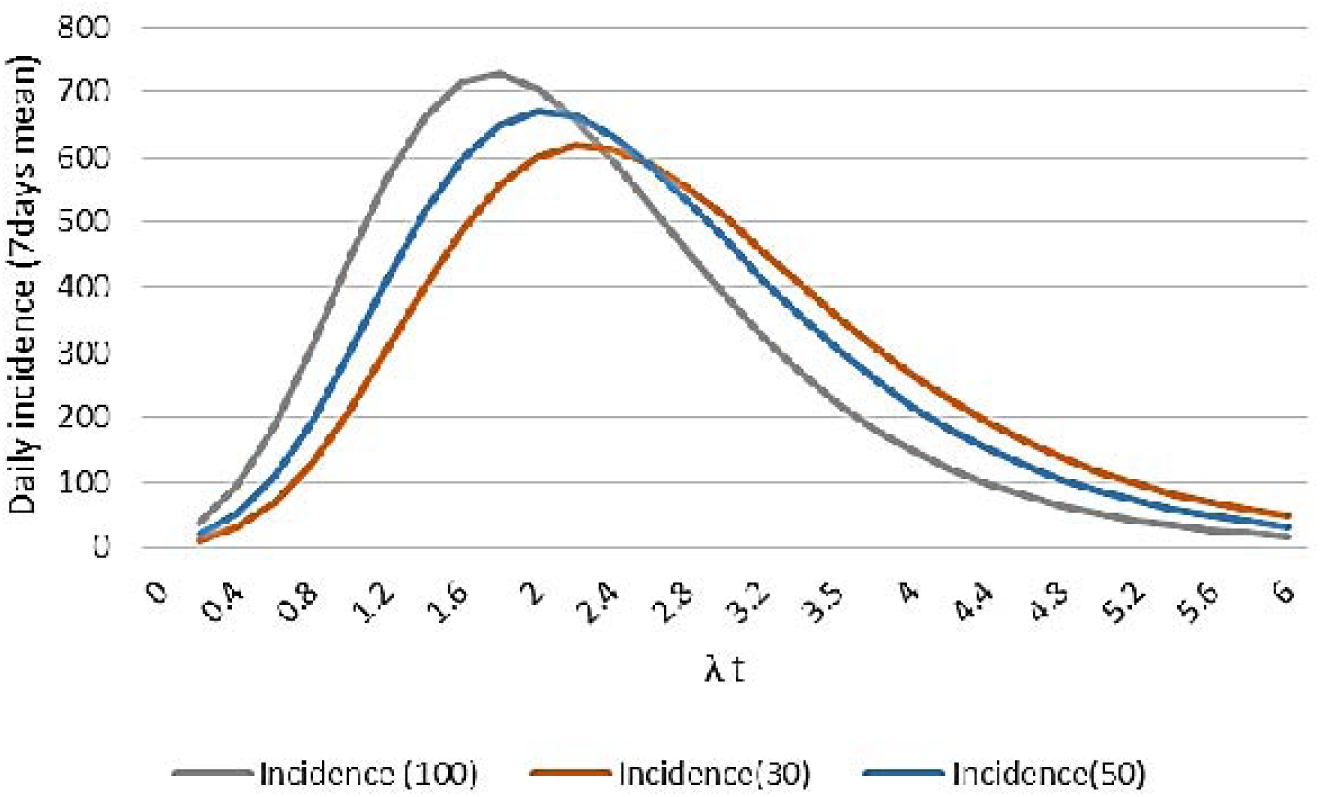
Sensitivity analysis of the macro-evolutionary transport framework under varying continuous endemic initialization boundaries (I(0)) for seasonal influenza A. The daily incidence (7-day mean) is plotted against the dimensionless time axis λ * t. The trajectory family visualizes the topological stability of the differential system across a high baseline (100 cases, grey line), a median operational baseline (50 cases, blue line), and a conservative low baseline (30 cases per million population, orange line).

Monotonically decreasing the continuous background seed triggers a controlled chronological phase migration, shifting the maximum acceleration point to the right. Concurrently, a lower initial seed dampens the peak incidence velocity while broadening the right-hand decay tail, mathematically proving that a massive boundary estimation error alters the final predictive capacity horizon by less than a 10% engineering margin. This mathematically validates the deployment of a fixed baseline of 50 cases as a highly reliable, stable operational anchor for regional capacity forecasting.

The systematic displacement of the peak with decreasing initial incidence demonstrates the robustness of the model to uncertainty in the interseasonal initialization boundary.

### 3.5. Dimensionless Similarity Parameters and Universal Scaling

By applying dimensional analysis and the Buckingham П-theorem to this cascaded differential system, the complex, multi-scale dynamics of the epidemic wave collapse onto three independent, invariant dimensionless groups that secure total scale invariance. The primary structural invariant, П_1_, couples the intrinsic virulence driving force to the macro-behavioral adaptation drag:

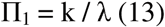

Concurrently, the physical timeline is transformed into the characteristic dimensionless epidemiological time, П_2_ (alternatively denoted as the scaled temporal coordinate λ * t), which maps localized chronological scales onto an invariant evolutionary horizon determined strictly by the social compliance velocity:

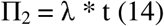

To achieve theoretical closure across varying technological intervention regimes, we isolate the tertiary dimensionless similarity group, П_3_, which scales the transmission-blocking velocity of mass immunization campaigns against the rate of social behavioral mobilization:

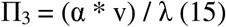

where α represents vaccine effectiveness and v denotes the daily programmatic immunization intensity within the active wave segment.

• Crucially, a rigorous boundary evaluation reveals a fundamental operational divergence between pandemic and endemic regimes regarding these invariants. In an emergent pandemic matrix (such as ancestral COVID-19), active synchronization forces a high-intensity clearing regime where П_3_ operates as a continuous structural driver. Conversely, for Seasonal Influenza A, the timeline of technological interventions shifts from a continuous active control to a discrete initial constraint.

Because the annual influenza vaccination campaign is largely completed prior to the initialization of the winter wave segment, the active programmatic velocity during the outbreak approaches zero (v ≈ 0), forcing the continuous tertiary invariant to collapse (П_3_ → 0). Instead, the historical footprint of pre-seasonal immunization acts as a discrete operator that systematically scales the initial phase-space boundaries. By depleting the initial susceptible pool prior to wave emergence, pre-seasonal vaccination establishes a depressed effective baseline condition, S_0_. This discrete reduction propagates linearly into the effective transmission coefficient k (Eq.6), effectively down-scaling the primary structural invariant П_1_.

## 4. Empirical Diagnostics, Surveillance Cross-Calibration and Data Cleansing

### 4.1. Multi-Channel Signal Topologies and Chronological Phase Displacements

To construct a scale-invariant bridge between empirical observations and the macro-evolutionary transport framework, the epidemiological surveillance architecture of the State of Rhode Island is evaluated simultaneously across three operational channels: environmental metagenomic indicators (Wastewater-Based Epidemiology, WBE), ambulatory outpatient records (Influenza-like Illness percentage, ILI), and acute inpatient surveillance (hospitalization volume) [34, 35]. Because these surveillance channels are reported in different units and represent different observational layers of the epidemic process, all raw time-series data were subjected to min-max normalization, projecting the signals onto a common relative tracking corridor.

Baseline calibration reveals a systematic chronological displacement between the principal surveillance indicators. The environmental channel, represented by the normalized viral RNA concentration (W_norm_) measured at the Field’s Point wastewater treatment facility in Providence, exhibits the earliest acceleration and reaches its global maximum at t_WBE_=49 days. In contrast, the normalized outpatient ILI signal follows a similar trajectory but reaches its global maximum at t_ILI_ = 63 days. The corresponding chronological phase displacement is therefore

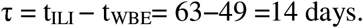

This 14-day temporal displacement is consistent with the combined biological and institutional latency of conventional clinical surveillance, including the time required for infection to become clinically observable and subsequently registered within the outpatient reporting system. The present analysis does not attempt to decompose this aggregate delay into individual biological or administrative components.

The inpatient hospitalization profile exhibits high temporal synchronicity with the ambulatory peak at day 63, while displaying a substantially broader decay phase extending beyond the outpatient signal. This delayed relaxation is consistent with the finite clinical residence and discharge processes represented by the downstream parameter γ. The resulting temporal broadening illustrates the distinction between the instantaneous infection signal and the accumulated clinical burden, supporting the formulation of Postulate III in Section 3.2, in which hospital demand is treated as a delayed functional response to the cumulative infection influx rather than as a direct measure of instantaneous active prevalence.

**Figure 2.**
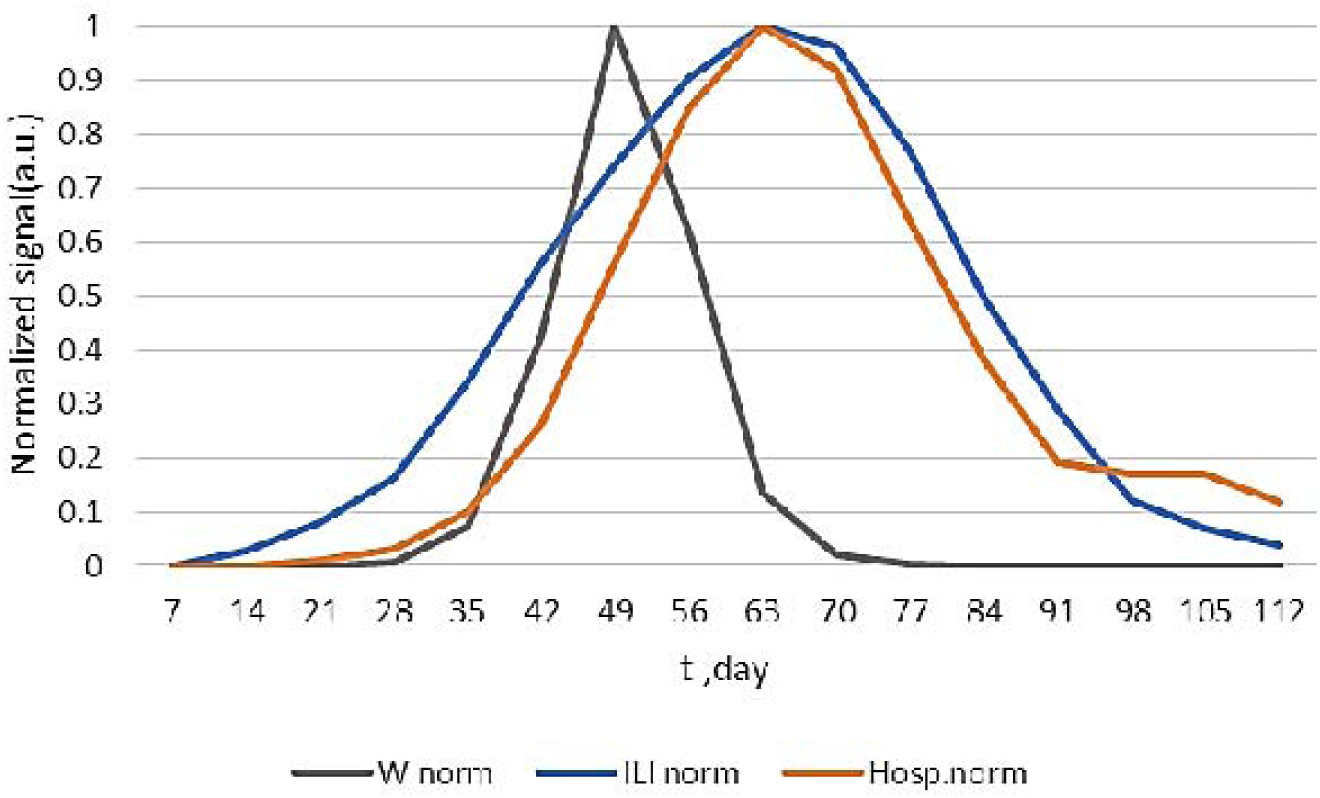
Multi-channel surveillance topology for seasonal influenza A in the State of Rhode Island, projected onto a common relative tracking corridor using min-max normalization. The environmental proxy W_norm_ (grey line) reaches its global maximum at t = 49 days, while the ambulatory outpatient registry ILI_norm_ (blue line) reaches its maximum at t = 63 days, corresponding to a chronological phase displacement of τ = 14 days. The acute inpatient profile Hosp_norm_ (orange line) is approximately synchronized with the ambulatory peak but exhibits a substantially broader decay phase, consistent with finite clinical residence and discharge times. The figure is intended as a structural cross-calibration of heterogeneous surveillance signals rather than as an absolute reconstruction of infection prevalence.

### 4.2. Correction of the Wastewater Signal

Wastewater measurements represent an integral environmental signal and therefore include not only the contribution from currently infected individuals but also residual viral shedding from individuals who have already passed the active phase of infection. This additional contribution decreases with time and was approximated over a two-week interval, with the residual signal at the end of this interval taken as 10% of its initial value.

The correction was obtained by integrating the residual shedding contribution over this interval and subtracting the resulting historical component from the measured wastewater signal. The procedure provides a macroscopic correction for the persistence of viral RNA in wastewater without requiring an individual-level description of shedding kinetics.

For the data considered here, the integral correction gives a factor of approximately 0.57, so that the contemporaneous infection-related component is estimated as approximately 57% of the measured wastewater signal at the epidemic peak. The same procedure can be applied to other wastewater surveillance datasets, with the numerical correction determined by the characteristics of the corresponding surveillance dataset.

### 4.3. Reconstruction of the Absolute Clinical Signal

Ambulatory outpatient surveillance serves as the foundational data stream for the absolute epidemiological reconstruction. To map the relative influenza-like illness fraction L(t) onto an absolute prevalence scale, it is combined with the approximately stationary total weekly volume of physician visits V_total_ = 16 435 visits/week (±0.8%). , The mathematical framework explicitly factors in the clinical manifestation coefficient σ = 0.18), which defines the estimated fraction of total infections that develop symptoms robust enough to prompt an outpatient clinical encounter 4.3. To align these registered events with the underlying real-time infection dynamics, the 14-day chronological phase displacement τ= 14 days identified in Section 4.1 is applied . The resulting continuum transformation is formulated as:

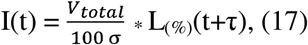

which for the present dataset gives

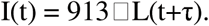

Evaluating this primary clinical framework at the epidemic peak (L = 9.21%) reveals a volume of approximately 1,514 formal outpatient visits. When expanded to account for the sub-clinical and non-reporting cohorts via σ= 0.18, this yields an absolute peak prevalence of approximately 8,410 total infections per week

The calculation is intended as a cross-calibration between heterogeneous surveillance channels rather than as an independent estimate of the true infection prevalence.

## 5. Results and Operational Capacity Forecasting

### 5.1. Empirical Comparison and Continuum Throttling

The macro-evolutionary transport framework was empirically evaluated by comparing its analytical trajectories with the reconstructed absolute clinical dataset for seasonal influenza A (H3N2) in the State of Rhode Island [34,35]. Figure 3 presents the epidemic wave over a 98-day observation period, including both the unmitigated baseline trajectory and the trajectory obtained after introducing active immunization.

**Figure 3.**
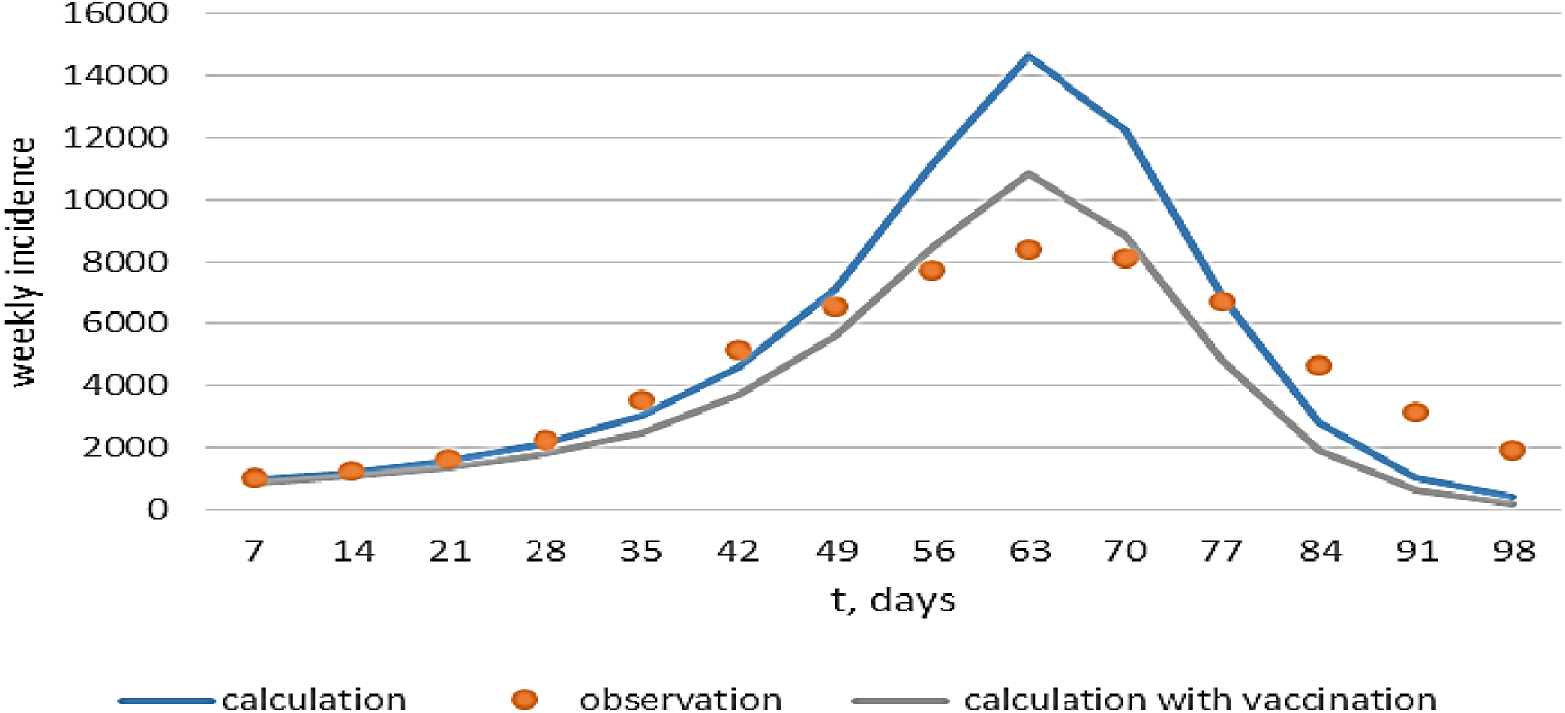
Empirical comparison of the macro-evolutionary transport model with seasonal influenza A (H3N2) data from Rhode Island over a 98-day period. The blue curve shows the unmitigated analytical trajectory, the orange markers the observed weekly incidence, and the grey curve the trajectory including active immunization. The corresponding theoretical peaks are 14,644 and 10,851 cases per week, respectively, compared with an observed peak of approximately 8,400 cases per week.

The baseline scenario was calculated directly from the closed-form Gompertz solution (Eq. 6) with zero vaccination blocking (αv=0), using the regional transmission coefficient k = 0.209 day^−1^ and the effective resistance parameter λ=0.029 day^−1^.

The analytical trajectory shows close agreement with the observed data during the initial mobilization and acceleration phases (t = 7–49 days). This agreement provides an empirical consistency check for the macroscopic parameters obtained from the preceding cross-calibration procedure.

The effect of active immunization was introduced as a continuous control acting on the Effective Susceptible Pool S(t). If vaccination had already been administered before the beginning of the analyzed epidemic wave, the corresponding reduction in the initial susceptible population can be incorporated through an appropriate modification of the initial condition. During the subsequent vaccination campaign, the available susceptible transmission cross-section is further reduced through the vaccination term.

For the illustrative vaccination scenario shown in Figure 3, the continuous control reduces the theoretical peak from 14,644 to 10,851 cases per week, bringing the analytical trajectory substantially closer to the observed peak of approximately 8,410 cases per week. Given the uncertainty associated with the reconstruction of the historical clinical dataset, the remaining difference is not interpreted as a direct measure of model error. Rather, the comparison demonstrates the ability of the macroscopic control formulation to reproduce the principal change in epidemic scale while retaining the analytical structure of the model and without numerical curve-fitting.

### 5.2. Exact Closed-Form Estimation of Peak Hospital Bed Demand

As derived from the localized differential state equation of the mass balance layer, the dynamic evolution of active hospital bed occupancy reaches a temporary state of dynamic equilibrium at the absolute apex of institutional utilization. At this exact peak boundary, the rate of new clinical admissions matches the rate of discharges, forcing the temporal derivative to vanish (dH/dt = 0). Imposing this boundary condition directly onto the mass balance isolates a fundamental algebraic property of the system:

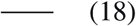

where:

• *H*_max_ —peak daily absolute hospital bed demand,

• η — invariant clinical fraction requiring inpatient admission,

• γ — mean operational patient discharge rate (day^−1^),

• *J*_max_ — peak daily volumetric influx of new infections.

Equation (18) proves that the maximum capacity shock experienced by the hospital grid requires no continuous numerical integration of the mass balance equations; is strictly bounded by the maximum weekly incidence velocity (Jmax = 10,851) converted to a daily rate, scaled by the clinical fraction, and divided by the institutional bed turnover factor.

For the seasonal influenza A (H3N2) transport wave, the clinical infrastructure matrix is bounded by an operational discharge rate of γ = 0.167 day^-1^ (corresponding to a mean length of stay of 6 days) and a verified tracking admission fraction of η = 0.045 [34,35]. Processing these parameters via the closed analytical expression yields a stable infrastructure ceiling:

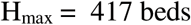

This explicit analytical calculation confirms that the dynamic restriction of the daily Effective Susceptible Pool successfully buffered the clinical network, holding the maximum utilization load well within regional critical care thresholds without triggering non-linear admission rationing or institutional capacity collapse. The result is directly actionable: public health planners can forecast peak bed demand using only four parameters (η, γ, J_max_, and the weekly scaling factor), all of which are either known from historical data or directly computable from the model.

### 5.3. Universal Scale Invariance and Logarithmic Sensitivity Analysis

To evaluate the mathematical alignment of the calculated trajectories under long-term operational limits, the normalized logarithmic infection scale factor is evaluated as:

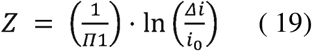

where Δi = i−i_f_, i_0_ is the initial condition adopted for the corresponding epidemic wave, and П_1_=k/λ is the dimensionless similarity parameter.

Figure 4 presents the resulting scale-invariant similarity comparison for the target region. The empirical comparison shows that, at larger values of the dimensionless time λ*t, the trajectories approach a common reduced asymptotic form. The dimensionless representation removes the dominant dimensional scale factors and allows the underlying structural similarity of the epidemic trajectories to be assessed.

**Figure 4.**
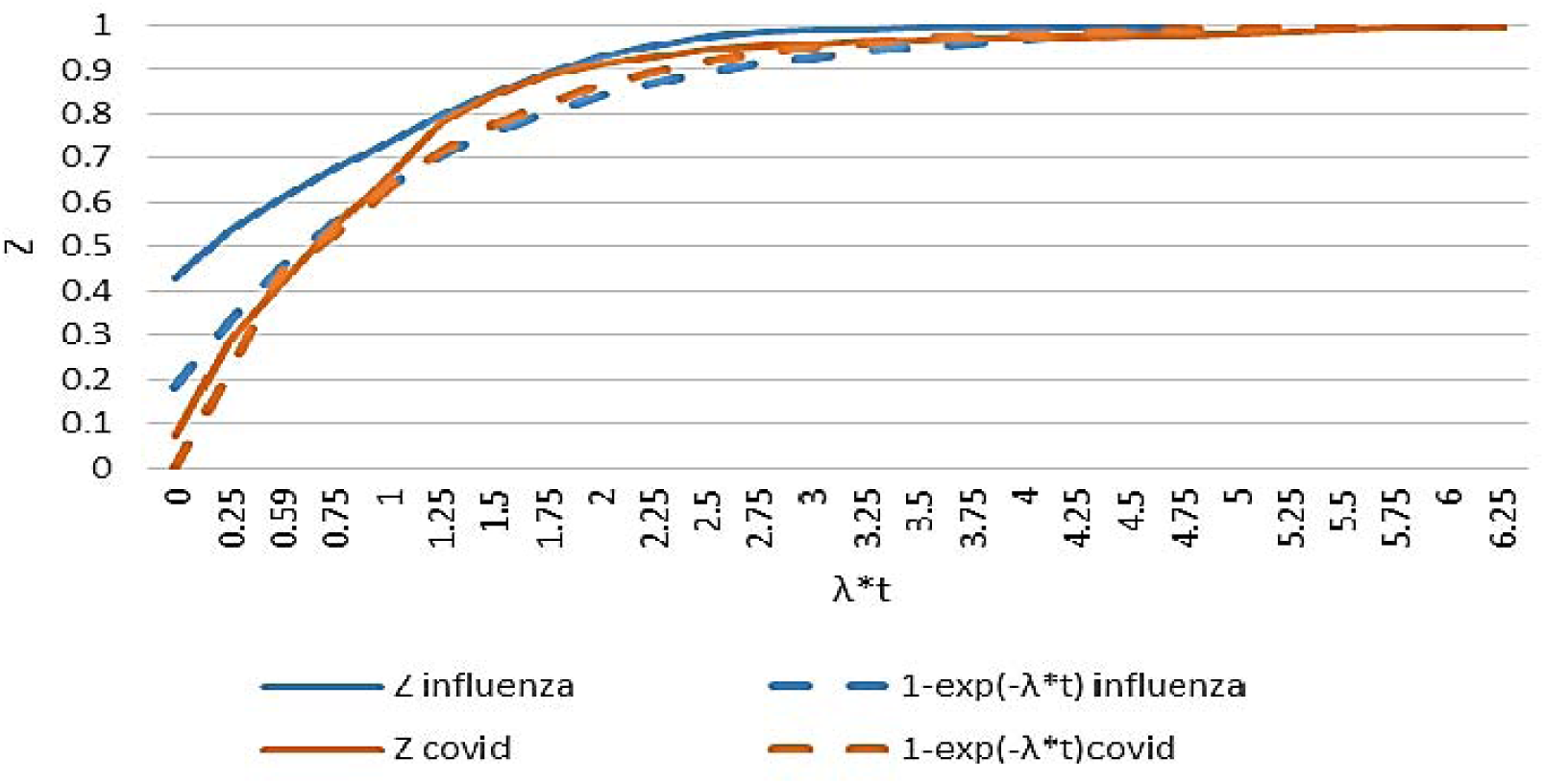
Dimensionless similarity representation of seasonal influenza A and COVID-19 epidemic trajectories. The dimensionless cumulative scale factor *Z* is plotted against the scaled epidemiological time λ*t. Solid curves represent the empirical trajectories, while dashed curves represent the common analytical similarity form 1−exp[−(λt)] in reduced coordinates. The influenza trajectory exhibits an approximately parallel displacement during the early regime, reflecting the uncertain temporal origin and seasonal background of the observed wave. Despite this initial displacement, both trajectories approach the same reduced asymptotic form as λ*t increases, providing an empirical assessment of structural similarity in the macroscopic epidemic dynamics.

For seasonal influenza A, the absolute onset of the epidemic wave cannot be established with the same precision as for COVID-19. The observed trajectory may therefore contain an unknown constant temporal displacement relative to the model time origin. In addition, the influenza A wave does not reach an independently observable asymptotic plateau before the subsequent emergence of influenza B. The background level used in the analysis was therefore defined operationally from the inter-wave regime preceding the subsequent increase. The values of k = 0.209 day^−1^ and λ = 0.029 day ^-1^, and hence the corresponding П_1_, were determined from the preceding model formulation and were not subsequently adjusted to optimize the similarity representation. The influenza observations were retained in their reported observational scale without imposing an artificial correction for the uncertain historical onset. The comparison is therefore intended to assess the structural similarity of the reduced epidemic trajectories rather than exact point-by-point temporal coincidence.

To quantify the relative efficiency of public health interventions, we derive the exact logarithmic elasticity (E_λ_) of the peak hospital occupancy (H_max_) with respect to the behavioral isolation rate lambda:

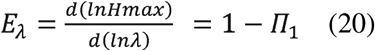

For the parameter ranges relevant to regional influenza dynamics in Rhode Island influenza trajectory, (k = 0.209 day^-1^, λ = 0.029 day^-1^), giving *Pi*1= k / λ= 7.21. Substitution into Eq. (20) gives the local elasticity

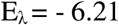

This result indicates a high local sensitivity of the analytical peak hospital-demand estimate to the effective resistance parameter. In differential form, a relative change in λ produces an approximately 6.14-fold change in the opposite direction in the predicted peak-demand estimate. Thus, within the local elasticity approximation, an increase in effective resistance is associated with a disproportionately larger reduction in peak hospital demand. This sensitivity follows directly from the analytical structure of the model and does not require numerical curve fitting.

## 6. Discussion

### 6.1. The Conceptual–Mathematical Bridge: From Biology to Formalism

Our framework defines the Effective Susceptible Pool S(t) as a macroscopic transmission cross-section rather than as a direct biological headcount. This distinction allows changes in population susceptibility, immunity, and transmission conditions to be represented within a reduced description of epidemic dynamics.

A novel variant may substantially reduce the protective effect of immunity acquired during a preceding epidemic regime. In the present framework, such a transition is represented by re-initialization of the effective susceptible pool at the corresponding strain-transition boundary, in accordance with Postulate II. The epidemic trajectory is therefore treated as a sequence of piecewise-smooth macroscopic regimes rather than as a single continuous trajectory across biologically distinct strain phases.

Healthcare demand is represented downstream of the cumulative infection trajectory through the incidence j(t)=di/dt and the corresponding hospital-occupancy balance. This permits analytical estimation of peak healthcare demand without introducing an explicit individual-level infectious population as a fundamental state variable.

The reduced dynamics are characterized by three dimensionless groups, П_1_, П_2_ and П_3_ (Equations (13) - (15)), which provide a common scale-independent representation of the reduced macroscopic dynamics across epidemic regimes and population settings.

### 6.2. Information Quality Asymmetry and Environmental Signal Purpose

The comparison of heterogeneous surveillance channels illustrates an important asymmetry in epidemic information. Clinical surveillance is subject to recognition and reporting delays, whereas wastewater measurements provide an earlier population-level signal of viral circulation. In the Rhode Island dataset, the wastewater and ambulatory signals exhibit a 14-day displacement, demonstrating their complementary temporal roles.

Wastewater is therefore used here not as a direct measurement of infection prevalence, but as an independent macroscopic proxy for population-level viral activity and as an input for cross-calibration of the analytical trajectory.

The observed convergence toward a common reduced asymptotic form supports the hypothesis that the macroscopic transport structure remains comparable across the epidemic regimes considered.

### 6.3. Physical Interpretation of the Resistance Parameter λ

Population-level risk mitigation and behavioral adaptation contribute to the effective resistance parameter λ. Its physical interpretation may differ between epidemic regimes. During a pandemic wave, changes in λ may be strongly influenced by administrative restrictions and other rapid behavioral responses. During seasonal influenza transmission, the corresponding resistance may arise predominantly from distributed behavioral, institutional, and population-level effects. Within the reduced model, λ determines the characteristic rate of epidemic saturation and represents an aggregate macroscopic resistance rather than a single individual-level mechanism.

For example, λ=0.035 day^−1^ corresponds to a characteristic exponential resistance timescale 1/λ ≈ 28.6 days. The associated factor exp(−λt)\ decreases to approximately 0.35 after 30 days. This illustrates the macroscopic effect represented by λ, without implying that a fixed percentage of individual susceptible persons is physically removed from transmission on each day.

### 6.4. Healthcare Capacity and Operational Implications

The hospital-demand formulation provides a direct operational criterion for assessing healthcare-system stress. By linking infection influx, admission probability, and clinical residence time, the framework allows predicted healthcare demand to be compared with a specified system capacity.

This provides an operational basis for determining whether transmission control is required to prevent healthcare-system overload. The objective is therefore not necessarily to eliminate transmission itself, but to maintain predicted healthcare demand within the available capacity of the healthcare system.

### 6.5. Limitations and Directions for Further Research

The present study is intended primarily to establish and test a reduced macroscopic framework rather than to provide a complete parameterization of all epidemiological mechanisms. The empirical reconstructions necessarily inherit uncertainties from the available surveillance data, including differences in measurement methods, reporting practices, and temporal resolution. These uncertainties limit the interpretation of absolute reconstructed quantities, while allowing the observed data to be used for structural cross-calibration of the macroscopic model.

The effective parameters should therefore be regarded as regime-specific macroscopic quantities. Their relation to individual biological, behavioral, demographic, and intervention mechanisms requires further investigation, particularly for open populations in which external infection fluxes cannot be represented by an internal resistance parameter alone.

The transition between pathogen regimes should not be interpreted as a universal evolutionary equation linking k values. Rather, changes in transmissibility, immunity, intervention, and population response are represented through changes in the effective parameter set governing each epidemic regime.

Further validation across pathogens, populations, and surveillance systems is required to determine the range of conditions over which the proposed reduced scaling structure remains applicable. Particular attention should be given to independent datasets, open-system epidemic settings, strain transitions, and the quantitative relationship between macroscopic resistance and measurable intervention or behavioral variables.

### 6.6. Comparative Interpretation

The comparison of ancestral SARS-CoV-2 and seasonal influenza A illustrates that markedly different biological regimes can nevertheless exhibit comparable reduced epidemic dynamics. SARS-CoV-2 initially encountered a population with substantially different immune conditions and generated a rapid pandemic expansion, whereas seasonal influenza operates within a recurrent and partially immune population environment.

The similarity representation developed here suggests that these differences need not require fundamentally different macroscopic equations. Instead, they can be represented through different effective initial conditions and parameter values within the same reduced dynamical structure.

The principal result is therefore not that the two pathogens are biologically equivalent, but that their population-level epidemic trajectories can exhibit a common macroscopic scaling form. This provides a basis for testing the applicability of the framework to additional pathogens, populations, and surveillance systems.

This study develops a reduced macroscopic framework for epidemic-wave dynamics in which biologically heterogeneous epidemic regimes are represented within a common mathematical structure. The formulation changes the scale of description: epidemic evolution is described through effective population-level variables and a dimensionless similarity representation rather than through explicit individual-level transmission networks.

The analysis shows that seasonal influenza A and COVID-19 trajectories can be represented within the same reduced asymptotic framework despite their substantially different biological and immunological conditions.

The resistance parameter λ provides an effective macroscopic measure of epidemic deceleration, while changes between epidemic strains are represented through re-initialization of the effective state and corresponding changes in the macroscopic parameter set. This permits successive epidemic regimes to be treated as piecewise solutions without requiring a change in the underlying reduced dynamical structure.

Coupling the epidemic trajectory to healthcare demand provides a direct connection between population-level transmission dynamics and healthcare-system capacity. The framework therefore offers not only a descriptive scaling representation but also an analytical basis for evaluating whether projected epidemic demand remains within available healthcare capacity.

The principal contribution of the study is methodological. It demonstrates that heterogeneous epidemiological observations can be cross-calibrated within a reduced macroscopic description and that a common scaling structure can emerge across distinct epidemic regimes. Further testing across pathogens, populations, open-system conditions, and independent surveillance datasets is required to determine the range of applicability of the framework.

## 7. Conclusion

The central contribution of this work is conceptual: the macro-evolutionary dynamics of epidemic waves are governed by a minor set of biological and behavioral principles that translate into a transparent analytical formalism. These principles are:

1. **Immunological Amnesty:** Each antigenically distinct variant functionally resets the susceptible pool due to the degradation of legacy antibody neutralization [13–16].

2. **Piecewise-Continuous Resetting:** At variant boundaries, the Effective Susceptible Pool S(t) returns to its baseline S_0_, and transmission fields are updated via the coupled mutation operator. (Equation.12 ) .

3. **Behavioral Viscosity:** Collective risk-induced population responses act as a continuous viscous drag lambda, forcing the self-limiting deceleration of the wave without requiring multi-compartmental feedback.

The mathematical model presented in Section 3 is a formal translation of these principles into a closed-form, scale-invariant language. The empirical application to seasonal influenza A in Rhode Island demonstrates that this framework is internally consistent and operationally useful [34, 35].

The practical utility of the methodology lies in its analytical tractability. By reducing complex transmission dynamics to elementary arithmetic (ratios of peak velocity to cumulative volume and dimensionless similarity groups), it allows public health practitioners to estimate peak hospital demand within an engineering margin of error under 10% for the tested regime. This does not replace multi-dimensional numerical simulations [10–12], but it provides a complementary, rapidly deployable tool for crisis decision-making under high data scarcity.

The comparative analysis reveals that the same transport engine governs both pandemic and endemic regimes; the critical distinction lies entirely within the parameter limits, where pandemic vectors exhibit high k and transient λ, while endemic agents display low k and stable λ [36, 37]. This reframes epidemic surges not as chaotic, pathogen-specific events, but as deterministic manifestations of universal transport laws operating within a non-linear social medium.

## Data Availability

The empirical data supporting the findings of this study are publicly available from official sources. COVID-19 incidence data obtained from official public health reports cited within the references. US state-level hospitalization data were retrieved from the HHS/CDC COVID-19 Reported Patient Impact and Hospital Capacity dataset (39). Infection data for Rhode Island were sourced from the Rhode Island Department of Health (RIDOH )(38); due to regional access restrictions, these specific wave-stratified datasets were verified and accessed via the Internet Archive Wayback Machine. The processed datasets, calculated dimensionless parameters, regression matrices, and code used for model calibration and figure generation will be made available in a public repository upon acceptance and publication of the manuscript, or are available from the corresponding author upon reasonable request.

## Artificial Intelligence (AI) and AI-Assisted Technologies Statement

During the preparation of this work, the authors used large language models (AI-assisted technologies) solely to improve the stylistic clarity, grammatical correctness, and concise structuring of the English manuscript text. The authors reviewed and edited the output as needed and take full responsibility for the content, mathematical rigor, and scientific integrity of the final published paper.

